# Hip shape shows a causal effect on hip fracture but not hip osteoarthritis: findings from a GWAS meta-analysis and causal analyses

**DOI:** 10.1101/2024.01.26.24301811

**Authors:** BG Faber, M Frysz, J Zheng, H Lin, KA Flynn, R Ebsim, FR Saunders, R Beynon, JS Gregory, RM Aspden, NC Harvey, C Lindner, T Cootes, D M. Evans, G Davey Smith, X Gao, S Wang, JP Kemp, JH Tobias

## Abstract

**Objectives:** Hip shape is thought to be an important causal risk factor for hip osteoarthritis and fracture. We aimed to identify genetic determinants of hip shape and use these to assess causal relationships with hip osteoarthritis.

**Methods:** Statistical hip shape modelling was used to derive 10 hip shape modes (HSMs) from DXA images in UK Biobank and Shanghai Changfeng cohorts (n_total_=43,485). Genome-wide association study meta-analyses were conducted for each HSM. Two-sample Mendelian randomisation (MR) was used to estimate causal effects between HSM and hip osteoarthritis using hip fracture as a positive control.

**Results:** Analysis of the first 10 HSMs identified 290 independent association signals (P<5×10^-8^). Hip shape SNPs were also associated (P<1.7×10^-4^) with hip osteoarthritis (n=29) and hip fracture (n=4). Fine mapping implicated *SMAD3* and *PLEC* as candidate genes that may be involved in the development of hip shape and hip osteoarthritis. MR analyses suggested there was no causal effect between any HSM and hip osteoarthritis, however there was evidence that HSM2 (higher neck-shaft angle) and HSM4 (wider femoral neck) have a causal effect on hip fracture (OR_IVW_ 1.27 [95% CI 1.12-1.44], P=1.79×10^-4^ and OR_IVW_ 0.74 [0.65-0.84], P=7.60×10^-6^ respectively)

**Conclusions:** We report the largest hip shape GWAS meta-analysis that identifies hundreds of novel loci, some of which are also associated with hip osteoarthritis and hip fracture. MR analyses suggest hip shape may not cause hip osteoarthritis but is implicated in hip fractures. Consequently, interventions aimed at modifying hip shape in older adults to prevent hip osteoarthritis may prove ineffective.

**Key messages:** *WHAT IS ALREADY KNOWN ON THIS TOPIC:* Hip shape in many forms has been linked with an increased risk of hip osteoarthritis and hip fracture. These observational associations have led to the inference of causality, prompting the development of surgical treatments aimed at modifying hip shape to potentially prevent hip osteoarthritis. Unfortunately, observational studies are susceptible to confounding and reverse causation.

*WHAT THIS STUDY ADDS:* This study provides a comprehensive catalogue of genetic associations related to variations in hip shape, in the form of 10 orthogonal hip shape modes. Substantial genetic overlap was observed between hip shape and both hip osteoarthritis and fracture. However, MR analyses suggested there was no causal effect between hip shape and hip osteoarthritis. Conversely, there was strong evidence that hip shape variation, including greater neck-shaft angle, is causal for hip fractures.

*HOW THIS STUDY MIGHT AFFECT RESEARCH, PRACTICE OR POLICY:* This study suggests that, at a population level, moderate hip shape variation does not cause hip osteoarthritis, meaning previously seen observational associations are likely confounded or due to reverse causality. Therefore, targeting these variations of hip shape through surgery, especially in older populations, may prove ineffective in preventing hip osteoarthritis.

## Introduction

Hip shape changes have been associated with hip osteoarthritis, with previous studies concluding that there is a causal link between the two (1, 2). However, observational studies are prone to confounding (3, 4). For example, it has been reported that young athletes are at increased risk of cam morphology, a bulging of the lateral aspect of the femoral head, which has been associated with hip osteoarthritis (1, 5, 6). However, this observed association might result from confounding due to excess physical activity causing hip osteoarthritis, rather than cam morphology. Differentiating between a causal and confounded association is important as surgical correction of hip shape has been proposed to prevent or delay the onset of hip osteoarthritis and trials are ongoing (7, 8); it has been difficult to test this hypothesis through randomised control trials due to the slow onset of hip osteoarthritis (3).

Mendelian randomisation (MR) uses genetic instruments obtained from genome-wide association studies (GWAS) to test for causal associations. This method is less susceptible to confounding and has been likened to a natural randomised control study that does not require years of follow up (9-11). We recently used MR to investigate whether cam morphology, as reflected by alpha-angle, has a causal effect on hip osteoarthritis prevalence, with null findings (12). However, it’s possible other aspects of hip shape have a causal effect on risk of hip osteoarthritis. Statistical shape modelling provides a holistic measure of hip shape by deriving principal components, also termed hip shape modes (HSMs), from points placed around the joint outline on images (13, 14). In a prior study using UK Biobank (UKB) data, 7 out of the 10 HSMs, which collectively explained the majority (>85%) of hip shape variance, were associated with both radiographic and hospital diagnosed hip osteoarthritis, as well as total hip replacement (15), suggesting these imaging phenotypes are clinically relevant. The automated approaches used to derive these HSMs from substantial numbers of participants provides an opportunity to conduct GWAS of multiple HSMs across different cohorts, which would in turn provide well powered genetic instruments that together represent a comprehensive proxy for hip shape, suitable for use in MR analyses.

Null findings in MR studies, such as our previous study of cam morphology, may reflect the lack of suitably powered genetic instruments. One way to assess the precision and power of genetic instruments for an exposure such as hip shape is to examine their relationships with another disease for which a causal effect is thought to exist. If a causal effect is found between the genetic instruments and one condition but not another, this makes it less likely the null findings result from inaccurate or under-powered genetic instruments. Hip fracture provides a good positive control for hip osteoarthritis given that measures of hip geometry, such as femoral neck width, are well established causal risk factors for hip fracture in both observational (16) and MR studies (insert Jon citation shortly).

In this study, we aimed to conduct a GWAS meta-analysis of the top 10 HSMs, in UKB and Shanghai Changfeng (SC) cohorts, where these HSM have been derived using our novel automated point placement method (17). Single nucleotide polymorphisms (SNPs) associated with hip shape were then used as genetic instruments in MR analyses evaluating whether hip shape is causally related to either hip osteoarthritis or hip fracture.

## Methods

### Hip shape measurement

In both UKB and SC (cohort details in Supplementary Methods), eighty-five landmark points were placed automatically around the proximal left femur to outline the femoral head, metaphysis, lesser and greater trochanters, and superior acetabulum. This automatic process has been described previously (15, 17). A statistical shape model was built from points fitted to all available images in UKB. Procrustes analyses removed size and rotational variation, producing a set of orthogonal principal components describing hip shape variation termed HSMs (15). The UKB statistical shape model was then applied to SC participants giving them scores for each HSM. The first 10 HSMs were selected for analyses, as together they explained the majority (86.3%) of hip shape variance in UKB (Supplementary Figure 1). HSMs and their associations with hip osteoarthritis have been described previously (15). Observational associations between HSMs and hip fracture were examined as part of this study (Supplementary Methods). The National Information Governance Board for Health and Social Care and Northwest Multi-Centre Research Ethics Committee (11/NW/0382) and UK Biobank Ethics Advisory committee gave ethical approval for all work in this study undertaken with UK Biobank data (UK Biobank application number 17295). The Zhongshan Hospital ethics committee affiliated to Fudan University gave ethical approval for this work (B2008-119(3)). All participants provided informed consent for this study.

### Hip shape genetic analyses

The first 10 HSMs (standardized to mean=0, standard deviation (SD)=1) were used as outcomes in GWAS adjusted for age, sex, genotyping chip and the first 20 ancestry principal components in UKB, and age, sex and the first 10 ancestry principal components in SC (see Supplementary Methods for genetic imputation and quality control). We tested SNP associations with each HSM assuming an additive allelic effect, in a linear mixed model implemented in BOLT-LMM v2.3.4 to account for cryptic population structure and relatedness in UKB. In SC we used fastGWA in GCTA v1.93.2 beta, a mixed linear model (MLM) approach to control for population stratification and relatedness. A centralized quality control of cohorts’ summary statistics was implemented in EasyQC (Supplementary Methods) (18). Both cohorts used the hg19 build. Given the trans-ancestry nature of our GWAS, an additive random effects meta-analysis was performed with METAL for each HSM (19, 20). Following meta-analysis, SNPs with MAF ≥0.01 in both cohorts were selected for further analyses. Conditional and joint genome-wide association analysis (GCTA-COJO) was used to identify statistically independent variants. Genome wide significant threshold was set at P<5×10^-8^. Figure 1 summarises the GWAS and fine-mapping methods (see Supplementary Methods for further details on GCTA-COJO, linkage disequilibrium score regression (LDSC) and fine mapping of loci).

**Figure 1.**
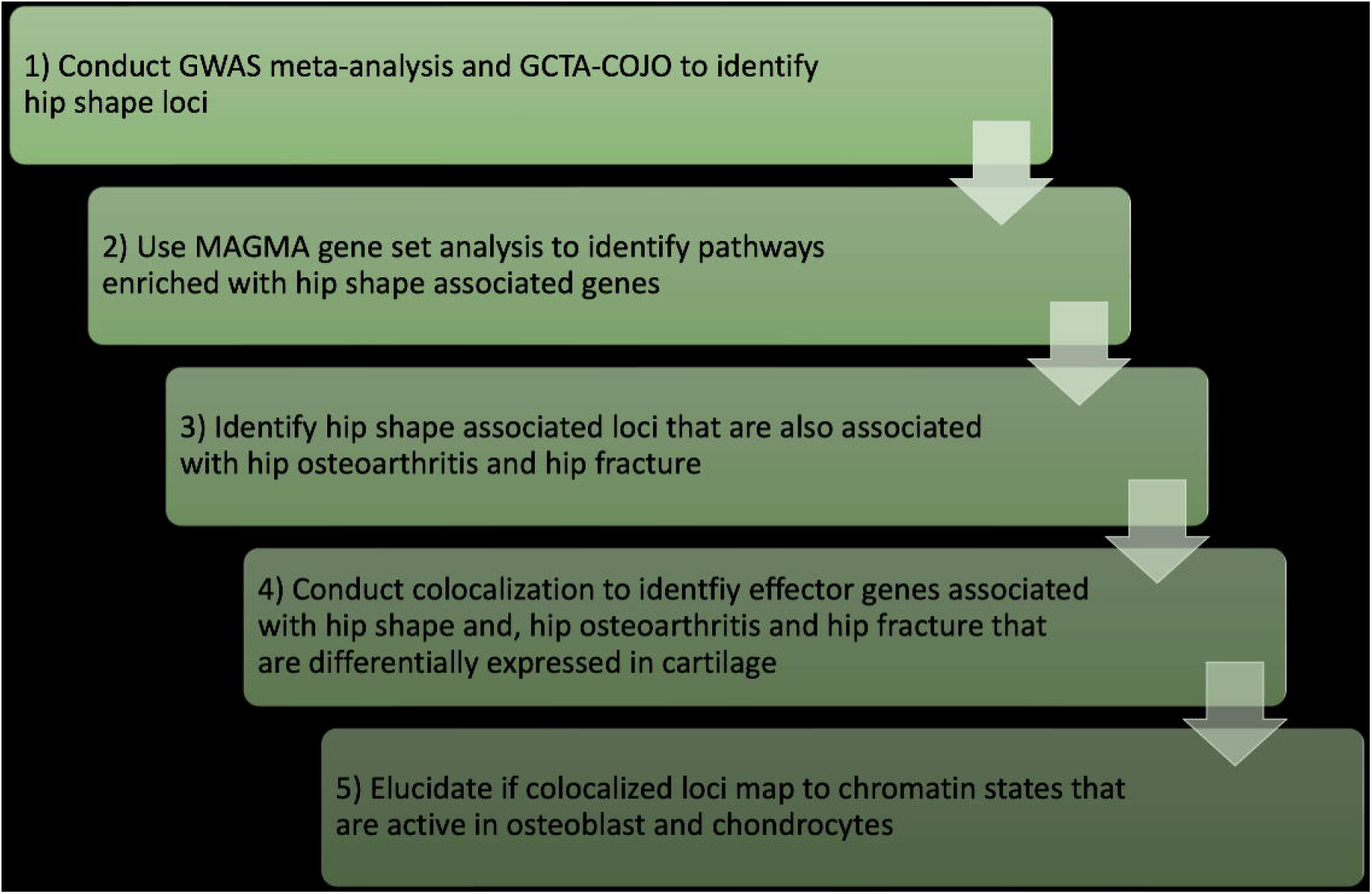
Five step process to identify genes involved with hip shape and hip osteoarthritis and hip fracture.

### Mendelian Randomisation

Two-sample MR, which uses genetic proxies as instrumental variables, was conducted to estimate the causal effect of each HSM on hip osteoarthritis and hip fracture using the TwoSampleMR R package (21). Conditionally independent variants from this study provided genetic instruments for HSMs. For outcomes, hip osteoarthritis data comprised a UKB GWAS of hospital diagnosed hip osteoarthritis, excluding participants included in the present HSM GWAS (12). Hip fracture data comprised a previous GWAS meta-analysis (22) which includes UKB (the 5% sample overlap with the hip shape GWAS is unlikely to bias the MR results (23)). The inverse-variance weighted (IVW) method was used as the primary analysis, where the causal estimate is obtained by combining the SNP-specific Wald ratios using a random-effects inverse-variance weighted meta-analysis. Strong evidence of a causal association was considered based on a Bonferroni adjusted P<0.005 (as 10 HSMs tested). Sensitivity analyses to test the robustness of our estimates, included weighted median and mode, simple mode and Egger regression (11). Reverse MR was applied to understand the causal effect of a genetic predisposition of hip osteoarthritis and hip fracture on HSMs. Effect estimates represent a one SD increase for continuous exposures (i.e. HSM) and a doubling of odds for binary exposures (i.e. hip osteoarthritis and fracture).

### Patient and Public Involvement

A local patient and public involvement group was consulted about study design and findings dissemination.

## Results

### Genome-wide Association Studies

For each HSM (1-10) (Figure 2) a GWAS meta-analysis was conducted in 43,485 participants (20,580 (47%)/22,905 (53%) male/female) across two studies (UKB n=38,175 & SC n=5,310) (see Supplementary Figures 2-11 for Manhattan plots). UKB participants were on average heavier and taller than the SC participants, but their ages were similar (Table 1). Furthermore, the mean HSM scores varied between the sexes and between UKB and SC (Table 1). A total of 298 independent associations at genome-wide significance (*P*<5.0×10^−8^) were found between SNPs and at least one HSM (after duplications were accounted for, where the same SNP was associated with different HSMs, 290 unique SNPs remained). These SNPs mapped to 182 loci of which 171 are novel signals, with 7 out of 9 previously identified HSM associated SNPs (24) remaining genome-wide significant (Figure 3, Supplementary Tables 1-11)). SNP heritability for the meta-analysed HSMs, as measured by LDSC, was between 14-21%, apart from HSM5, which had a lower SNP heritability of 6% (Table 2). SNP heritability estimates within each cohort, calculated with LDSC, were broadly similar (see Supplementary Table 12). Genomic inflation of the HSM GWAS was low (λ=0.86-0.94) likely due to the conservative random-effects model used (Supplementary Table 12). 15 SNPs showed at least suggestive evidence of colocalisation with mRNA expression in human joint tissue (Supplementary Table 13).

**Figure 2.**
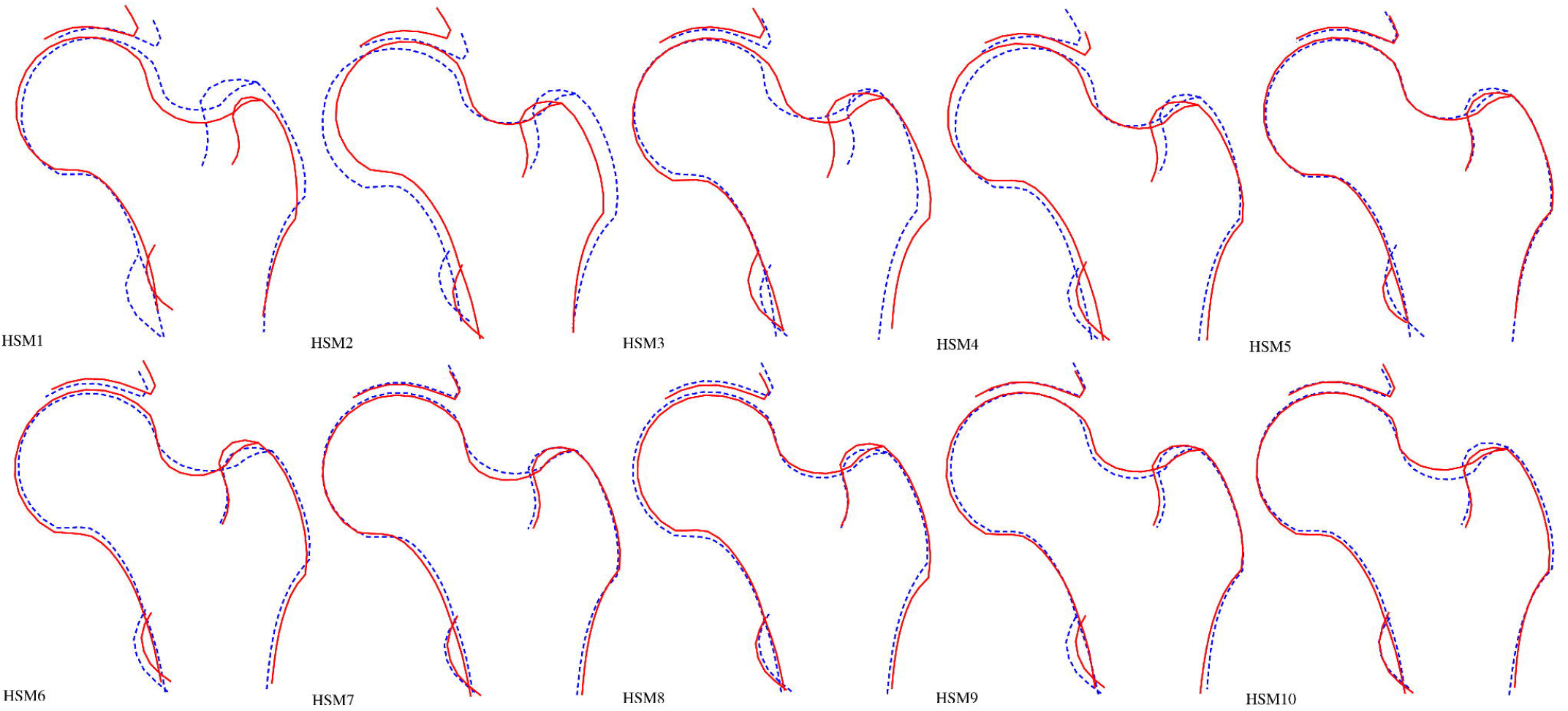
Visual representation of the first 10 hip shape modes. Each hip shape mode (HSM) represents statistically independent (orthogonal) variation in hip shape captured by hip DXA. HSM1 explains the most variation and each HSM there after explains less variation. The HSMs are plotted here where the dotted line represents the hip shape comprised of-2 standard deviation variation away from the mean and solid line represents +2 standard deviation hip shape away from the mean. Each participant in this study is given a score for each HSM which represents where their hip shape lies within each mode.

**Figure 3.**
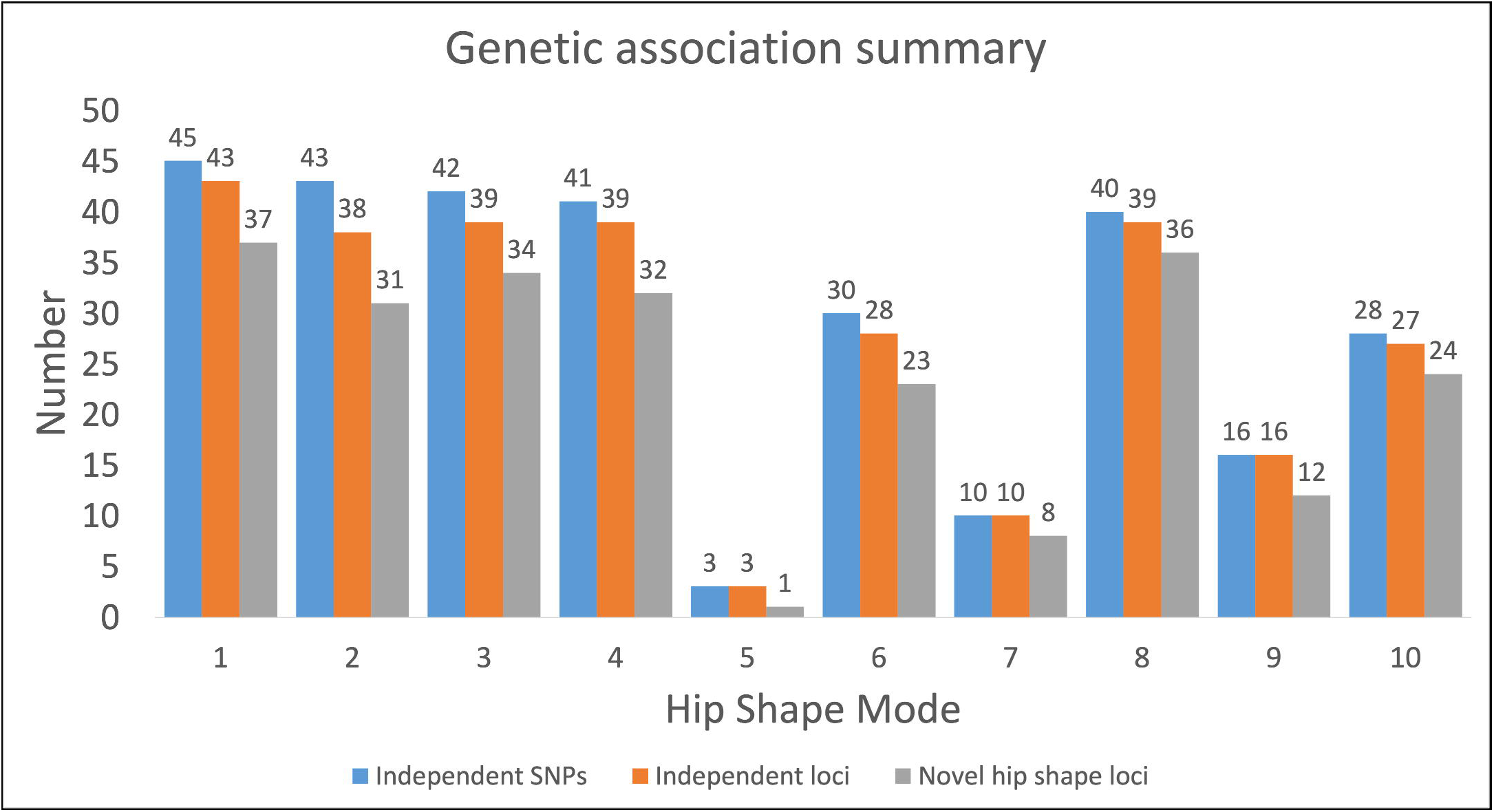
Genetic associations for each hip shape mode. Independent single nucleotide polymorphisms (SNPs) are those that are conditionally independent as obtained by GCTA-COJO. Independent loci are defined by the closest gene. Novel hip shape loci are those further than 1Mb from previously published SNPs associated with hip shape.

**Table 1.** Descriptive characteristics of study participants, combined and stratified by sex.

|  | UK Biobank |  |  | Shanghai Changfeng |  |  |
| --- | --- | --- | --- | --- | --- | --- |
|  | Male<br>Mean (range, SD) | Female<br>Mean (range, SD) | Combined<br>Mean (range, SD) | Male<br>Mean (range, SD) | Female<br>Mean (range, SD) | Combined<br>Mean (range, SD) |
| Age | 64.5 (45-81, 7.6) | 63.1 (45-82, 7.4) | 63.8 (45-82, 7.5) | 64.5 (45-81, 9.5) | 62.6 (46-88, 9.2) | 63.4 (46-96, 9.4) |
| Height | 177.3 (150-204, 6.6) | 163.7 (135-196, 6.4) | 170.2 (135-204, 9.4) | 168.0 (143-189, 6.1) | 156.8 (132-185, 5.9) | 161.6 (132-189, 8.2) |
| Weight | 83.3 (47-171, 13.4) | 68.2 (34-169, 12.8) | 75.4 (34-171, 15.1) | 69.2 (39-107, 9.8) | 59.2 (35-106, 9.1) | 63.5 (35-107, 10.6) |
| HSM1 | -0.31 (-4.56-3.57, 1.0) | 0.28 (-3.78-3.88, 0.9) | 0.00 (-4.56-3.88, 1.0) | -0.85 (-3.80-2.22, 0.94) | -0.36 (-4.15-2.70, 0.96) | -0.57 (-4.15-2.70, 0.98) |
| HSM2 | 0.02 (-4.53-4.48, 1.0) | -0.01 (-4.69-4.19, 1.0) | 0.00 (-4.69-4.48, 1.0) | -0.37 (-4.16-3.95, 1.01) | -0.47 (-4.96-3.30, 1.11) | -0.43 (-4.96-3.95, 1.07) |
| HSM3 | 0.32 (-3.62-4.26, 1.0) | -0.30 (-4.10-4.00, 0.9) | 0.00 (-4.10-4.26, 1.0) | 1.13 (-2.28-4.01, 0.94) | 0.83 (-2.45-4.05, 0.95) | 0.96 (-2.45-4.05, 0.96) |
| HSM4 | 0.14 (-3.83-3.99, 1.0) | -0.13 (-4.38-3.99, 1.0) | 0.00 (-4.38-3.99, 1.0) | 0.15 (-2.90-4.36, 0.92) | -0.003 (-3.07-3.96, 0.98) | 0.06 (-3.07-4.36, 0.96) |
| HSM5 | -0.41 (-4.52-4.38, 0.9) | 0.04 (-4.24-3.42, 1.1) | 0.00 (-4.52-4.38, 1.0) | -0.46 (-3.62-2.63, 0.91) | -0.02 (-3.44-2.92, 0.92) | -0.21 (-3.62-2.92, 0.94) |
| HSM6 | -0.26 (-4.58-3.88, 1.0) | 0.22 (-3.39-4.99, 1.0) | 0.00 (-4.58-4.99, 1.0) | 0.36 (-2.51-3.46, 0.89) | 0.59 (-2.36-3.57, 0.89) | 0.49 (-2.51-3.57, 0.90) |
| HSM7 | -0.10 (-4.58-5.05, 1.0) | 0.08 (-4.93-4.74, 1.0) | 0.00 (-4.93-5.05, 1.0) | -0.23 (-4.90-2.67, 0.91) | 0.17 (-2.56-4.03, 0.91) | 0.00 (-4.90-4.03, 0.93) |
| HSM8 | 0.08 (-4.84-4.55, 1.0) | -0.07 (-4.35-3.96, 1.0) | 0.00 (-4.84-4.55, 1.0) | -0.02 (-3.70-3.03, 1.00) | -0.17 (-3.47-3.57, 1.02) | -0.11 (-3.70-3.57, 1.01) |
| HSM9 | 0.31 (-3.65-5.03, 1.0) | -0.28 (-4.13-4.54, 0.9) | 0.00 (-4.13-5.03, 1.0) | 0.24 (-3.16-3.62, 0.96) | -0.14 (-3.46-3.10, 0.97) | 0.02 (-3.46-3.62, 0.99) |
| HSM10 | -0.01 (-4.09-3.84, 1.0) | 0.01 (-4.12-3.85, 1.0) | 0.00 (-4.12-3.85, 1.0) | 0.22 (-3.58-3.60, 1.01) | -0.08 (-4.28-3.41, 0.97) | 0.05 (-4.28-3.60, 1.00) |
| **N** | 18,646 | 20,374 | 38,175 | 2,263 | 3,047 | 5,310 |

Units are as follows: Age – years, Height – centimetres, weight – kilograms, hip shape modes (HSM) – standard deviations (SDs)

**Table 2.**
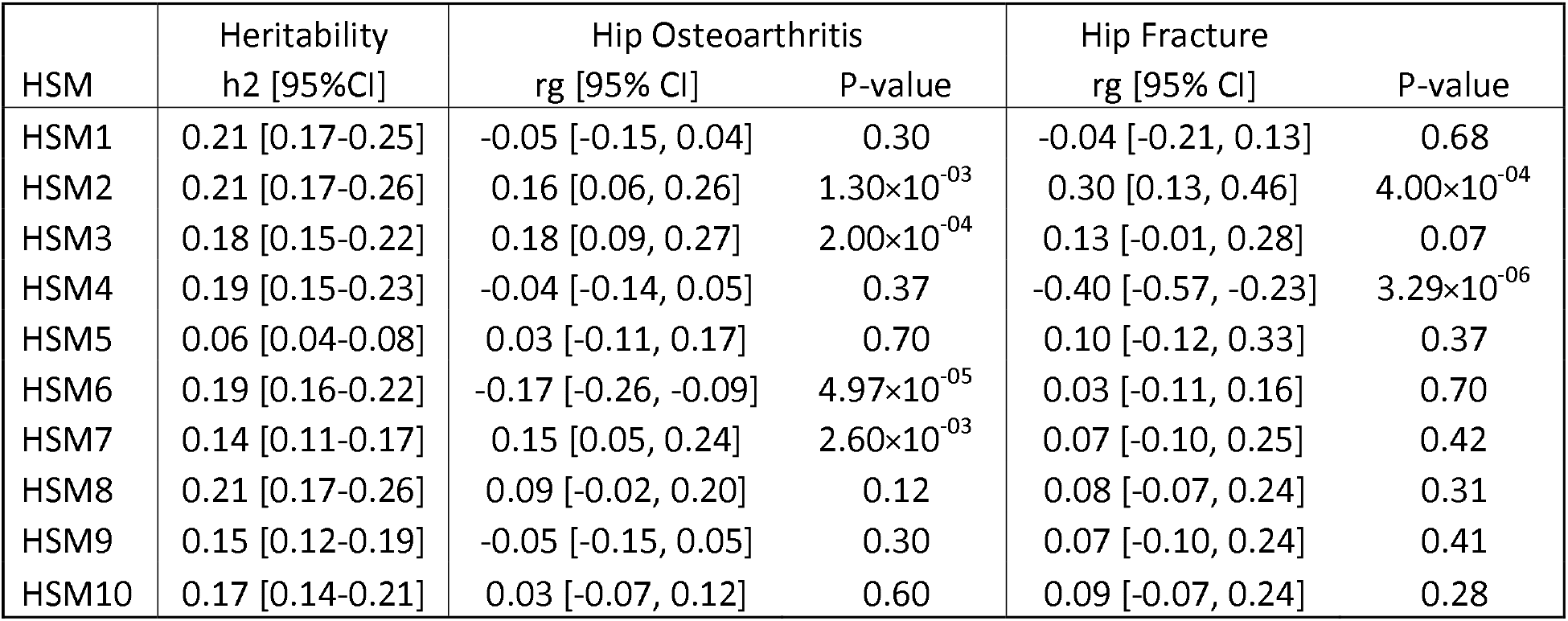
SNP heritability of the hip shape mode meta-analyses and their genetic correlations with hip osteoarthritis and hip fracture.

### Gene Set Analysis

To prioritise genes contributing to each HSM we applied MAGMA gene set analyses using a Bonferroni adjusted p-value threshold (P<2.70×10^-6^, 0.05/18824 number of gene sets tested). In total, 30 gene sets were associated with the HSMs (Supplementary Tables 14). Two gene sets showed associations with more than one HSM; the *skeletal morphogenesis* set was associated with HSM1 (Genes n=217 & P 2.98×10^-8^), HSM6 (Genes n=217 & P 3.98×10^-7^) and HSM10 (Genes n=217 & P 7.83×10^-8^), and the *response to growth factor* set was associated with HSM4 (Genes n=662 & P 1.27×10^-7^) and HSM10 (Genes n=662 & P 7.83×10^-8^). Seven gene sets were related to skeletal, bone and connective tissue development, 5 to cartilage formation, 4 to signalling pathways known to affect cartilage (TGF-β and growth factor). Details of the gene sets can be found in Supplementary Table 15.

### Genetic Correlation

LDSC was used to calculate the genetic correlation between HSM and different disease traits. For hip osteoarthritis, HSM2 (rg 0.16 [95% CI 0.06, 0.26]), HSM3 (0.18 [0.09, 0.27]), HSM6 (-0.17 [-0.26, -0.09]) and HSM7 (0.15 [0.05, 0.24]) showed some evidence of genetic correlation. For hip fracture, HSM2 (0.30 [0.13, 0.46]) and HSM4 (-0.40 [-0.57, -0.23]) showed a moderate genetic correlation (Table 2). The genetic correlations between UKB and SC, and with disease outcomes were assessed for each HSM but in SC were underpowered due to the smaller sample size of SC (Supplementary Table 16). Despite the HSMs being orthogonal there was weak to moderate genetic correlation between them suggesting shared underlying genetic aetiology (Supplementary Figure 12).

### Genetic overlap between hip shape and hip osteoarthritis

Conditionally independent HSM-associated SNPs were looked up in a previous GWAS of hip osteoarthritis (25). 29 SNPs were associated with a HSM (genome wide significance P<5×10^-8^) and hip osteoarthritis (Bonferroni adjusted P<1.7×10^-4^, to account for the 290 SNPs tested) (Table 3). 19 SNPs were >1Mb away from previously implicated hip shape loci and represented novel signals. To understand if the hip shape and osteoarthritis GWAS signals were shared Bayesian colocalisation was conducted which found 18 SNPs colocalised (PP >80%). In particular, two SNPs showed strong SNP-gene evidence after fine-mapping.

**Table 3.**
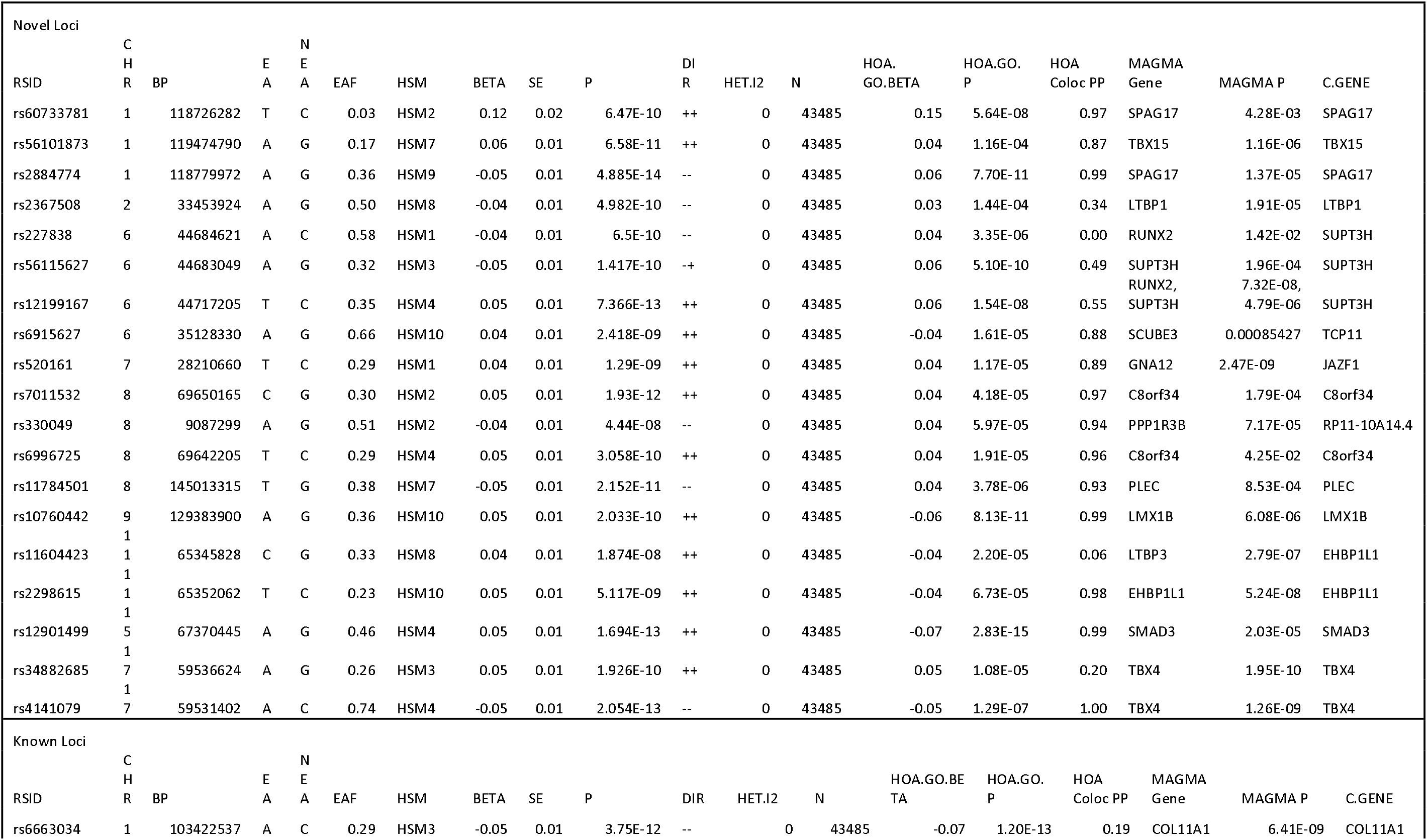
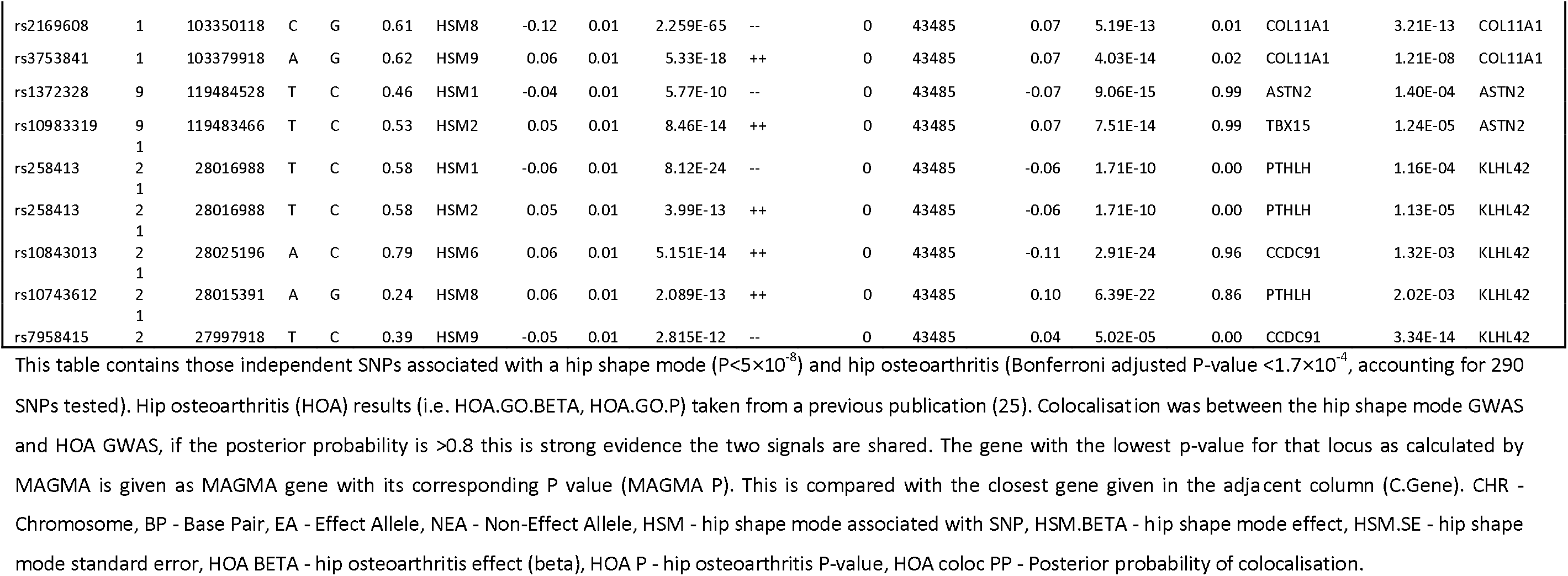
Shared hip shape and hip osteoarthritis signals.

Rs12901499, within the *SMAD3* locus, was positively associated with HSM4 (β 0.05, P 1.69×10^-13^), describing greater acetabular coverage and narrower femoral neck, and was also found to be protective of hip osteoarthritis (β -0.07, P 2.83×10^-15^). In addition, these two GWAS signals colocalised with each other (PP 99%) and the HSM4 GWAS signal colocalised with *SMAD3* mRNA expression in both healthy and degraded human cartilage (PP 98% in both tissues, Supplementary Table 13).

Rs11784501, within the *PLEC* locus, was negatively associated with HSM7 (β -0.05, P 2.15×10^-11^), describing a wider femoral neck, and was also found to increase the risk of hip osteoarthritis (β 0.04, P 3.78×10^-6^). Again, these two GWAS signals colocalised (PP 93%) and the HSM7 GWAS signal colocalised with *PLEC* mRNA expression in human synovial tissue (PP 87%, Supplementary Table 13). Rs11784501 was predicted to have a high chromatin state activity in both chondrocytes (chromatin state=2, encodes a region flanking a transcription starting site, 1 is the highest activity state and 15 is the lowest) and osteoblasts (chromatin state=1, encodes a variant at a transcription starting site) suggesting a role in altered transcription (Supplementary Table 17).

### Overlap between hip shape and hip Fracture

Despite low number of hip fractures (n=81), in models adjusted for age, sex, height and weight, two HSMs showed weak evidence of an observational association with hip fracture. HSM2, describing an increased femoral neck shaft angle and less acetabular coverage, was associated with an increased risk of hip fracture (odds ratio (OR) 1.26 [95% CI 1.01-1.56], P 0.04). HSM4, describing a narrower femoral neck and increased acetabular coverage, was protective of hip fracture (OR 0.78 [0.63-0.98], P 0.03) (Supplementary Table 18).

Four HSM-associated SNPs (P<5×10^-8^) also showed an association with hip fracture (P<1.7×10^-4^, to account for the 290 SNPs tested) (Table 4). Rs12475479 was negatively associated with HSM2 (β - 0.06, P 4.96×10^-14^), describing a reduced femoral neck-shaft angle and greater acetabular coverage, and was found to be protective of hip fracture (β -0.08, P 4.79×10^-5^). *DIS3L2* was implicated by MAGMA at this locus although colocalisation analyses showed little evidence of shared HSM2 and hip fracture GWAS signals. Three SNPs (rs56368105, rs736825 & rs11614913) on chromosome 12 and in close LD with each other (D’ 0.78-0.98, R^2^ 0.51-0.89) showed positive associations with HSM4 (a narrower femoral neck and larger femoral head, β 0.06, P 1.60×10^-18^), HSM9 (wider femoral neck and larger femoral head, β 0.08, P 4.73×10^-31^) and HSM10 (wider femoral neck, β 0.05, P 1.18×10^-14^). Rs56368105 (β -0.08, P 1.38×10^-7^) and rs736825 (β -0.08, P 8.87×10^-8^) were protective for hip fracture, whereas rs11614913 was associated with increased risk (β 0.08, P 4.73×10^-31^). All three SNPs were predicted to increase transcription in both chondrocytes (chromatin state 1, 2 and 2 respectively) with only rs56368105 increasing transcription in osteoblasts (chromatin state 1) (Supplementary Table 17). MAGMA implicated several genes at this locus including *HOXC9, RP11-834C11.12, HOXC8, HOXC5, HOXC6* and *HOXC4* but none of the signals colocalised with mRNA expression in human tissue.

**Table 4.** Shared hip shape and hip fracture signals.

| RSID | CHR | BP | EA | NEA | EAf | HSM | BETA | SE | P | DIR | HET.12 | N | HipFrac.Beta | HipFrac.P | Hip<br>Frac<br>Coloc<br>PP | MAGMA Gene | MAGMA P | C.GENE |
| --- | --- | --- | --- | --- | --- | --- | --- | --- | --- | --- | --- | --- | --- | --- | --- | --- | --- | --- |
| rs12475479 | 2 | 233041502 | A | T | 0.26 | HSM2 | -0.06 | 0.01 | 4.96E-14 | -- | 0 | 43485 | -0.08 | 4.79E-05 | 0.09 | DIS3L2 | 2.87E-06 | DIS3L2 |
| rs56368105 | 12 | 54393770 | A | G | 0.56 | HSM4 | 0.06 | 0.01 | 1.6E-18 | ++ | 0 | 43485 | -0.08 | 1.38E-07 | 0.10 | RP11-834C11.12, HOXC6 | 2.22E-16, 1.09E-14 | RP11-834C11.12 |
| rs736825 | 12 | 54417576 | C | G | 0.60 | HSM9 | 0.08 | 0.01 | 4.37E-31 | ++ | 0 | 43485 | -0.08 | 8.87E-08 | 0.08 | HOXC9, RP11-834C11.12, HOXC8, HOXC5, HOXC6 | 5.83E-15, 1.21E-13, 3.79E-11, 1.79E-10, 5.00E-10 | RP11-834C11.12 |
| rs11614913 | 12 | 54385599 | T | C | 0.43 | HSM10 | 0.05 | 0.01 | 1.18E-14 | ++ | 0 | 43485 | 0.08 | 1.30E-07 | 0.10 | HOXC9, HOXC6, RP11-834C11.12, HOXC8, HOXC4 | 1.76E-12, 7.25E-11, 3.39E-10, 7.27E-10, 1.60E-06 | RP11-834C11.12 |

This table contains those independent SNPs associated with a hip shape mode ( $P < 5 \times 10^{-8}$ ) and hip fracture (Bonferroni adjusted P-value $< 1.7 \times 10^{-4}$ ). Hip fracture (HipFrac) results taken from a previous publication (22). Colocalisation was between the hip shape mode GWAS and HipFrac GWAS, if the posterior probability is $> 0.8$ this is strong evidence the two signals are shared. CHR - Chromosome, BP - Base Pair, EA - Effect Allele, NEA - Non Effect Allele, HSM - hip shape mode associated with SNP, HSM.BETA - hip shape mode effect, HSM.SE - hip shape mode standard error, HipFrac.BETA - hip fracture effect (beta), HipFrac.P - hip fracture P-value, Hip.Frac coloc PP - Posterior probability of colocalisation.

### Mendelian Randomisation

MR analyses suggested there was no causal effect of any HSM on hip osteoarthritis (Figure 4, Supplementary Table 19). The mean F-statistics for the genetic instruments of the HSMs ranged from 37 to 56 (Supplementary Tables 1-10) indicating acceptable instrument strength (11). Reverse MR used 27 genetic instruments for hip osteoarthritis, which had a mean F-statistic of 45 (range 30-100), indicating acceptable instrument strength (Supplementary Table 20). Genetic predisposition to hip osteoarthritis was suggested to causally effect HSM3 (cam-type femoral head with bulging of the lateral aspect) (IVW β 1.37 [95% CI 0.54-2.20], P 1.21×10^-3^) (Figure 4, Supplementary Table 19). Sensitivity analyses showed the same direction of effect but with weaker statistical evidence (Supplementary Table 19).

**Figure 4.**
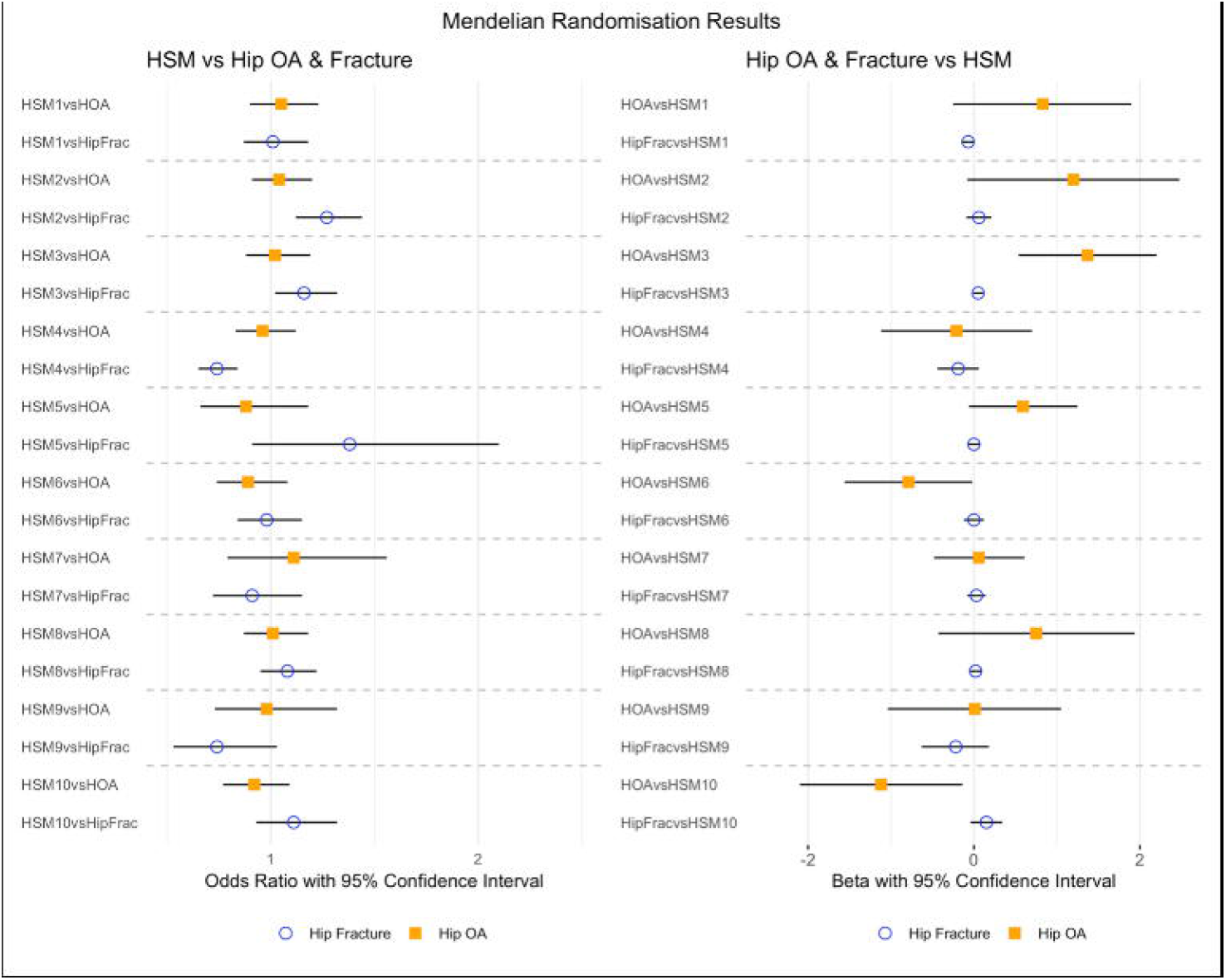
Bi-directional causal analyses between hip shape, and hip osteoarthritis and hip fracture.

Counter to the results seen for hip osteoarthritis, MR analyses showed strong evidence for HSM2 (higher neck-shaft angle and less acetabular coverage, as represented by the +2 SD line (Figure 1)) and HSM4 (less acetabular coverage and wider femoral neck, as represented by the -2 standard deviation (SD) line (Figure 2)) having a causal effect on hip fracture (IVW OR 1.27 [95% CI 1.12-1.44], P 1.79×10^-4^ and OR 0.74 [0.65-0.84], P 7.60×10^-6^ respectively) (Figure 4, Supplementary Table 21). These effects were broadly supported by the sensitivity analyses (Supplementary Table 21). There was weaker evidence that HSM3 (a cam-type femoral head and shorter femoral neck) may also have a causal effect on hip fracture (IVW OR 1.16 [95% CI 1.02-1.32], P 0.02] but the sensitivity analyses showed little evidence to support this. There was little evidence that a genetic predisposition to hip fracture affects any of the HSM modes (Figure 4, Supplementary Table 21). In addition, the three instruments for hip fracture derived from a previous meta-analysis had acceptable strength (F-statistic range 36-44) (Supplementary Table 22).

## Discussion

In this study we present the results from a GWAS of 10 orthogonal HSMs that identified 290 conditionally independent SNPs, mapping to 182 loci which is a marked increase on previous hip shape GWAS (24, 26). In gene set analyses, skeletal, bone and cartilage related pathways showed the strongest evidence of association with hip shape variance. HSMs were genetically correlated with hip osteoarthritis and hip fracture indicating shared underlying genetic aetiology. Of hip shape associated SNPs, 29 were also associated with hip osteoarthritis and 4 with hip fracture. MR analyses failed to show evidence of a causal effect of hip shape on hip osteoarthritis but did show a causal effect of hip shape, in the form of a higher neck shaft angle and reduced acetabular coverage, on hip fracture. In addition, there was evidence that a genetic predisposition to osteoarthritis caused a cam-type femoral head to develop.

Understanding the genetic aetiology of hip shape is important given its implication in the aetiology of hip osteoarthritis and hip fracture in observational studies (13-15, 27, 28). When investigating the overlap between hip shape loci and both conditions, there were many more hip shape SNPs associated with hip osteoarthritis than hip fracture (29 vs 4). This suggests there is a greater shared genetic aetiology between hip shape and hip osteoarthritis than with hip fracture. These results might also be expected given the increased genetic heritability of hip osteoarthritis (h^2^ 0.04) compared with hip fracture (h^2^ 0.004). Hip fractures by their nature are normally a consequence of falls (∼90%) that themselves are often related to environmental factors (29). In terms of specific genes likely to affect both hip shape and either hip osteoarthritis or fracture, *SMAD3* and *PLEC* were the only genes identified through colocalisation between GWAS signals and mRNA expression in human joint tissue and both were associated with hip osteoarthritis. *SMAD3* is an important member of the TGF-β pathway which plays an important role in chondrogenesis and has previously been implicated in hip osteoarthritis but was not known to be associated with hip shape (30, 31). *PLEC* encodes Plectin a large cytoskeleton protein that is important in regulating cellular responses to mechanical stressors and is thought to play a role in the development of osteoarthritis but has not previously been associated with hip shape (32).

The independent SNPs associated with each HSM were used as genetic proxies for the described hip shape variation in Mendelian randomisation, to estimate the causal effect of each HSM on hip osteoarthritis and fracture. There was little evidence that any HSM had a causal effect on hip osteoarthritis despite a previous study using the same participants in UKB showing strong observational associations between these HSMs and hip osteoarthritis and total hip replacement (15). These results counter the hypothesis that moderate alterations in hip shape (e.g. cam morphology or acetabular dysplasia) cause hip osteoarthritis in the general population (2, 7).

However, they align with our recent study, in the same participants, which found no strong evidence of a causal association between cam morphology (based on alpha-angle derived from the same points used to generate HSMs in the present study) and hip osteoarthritis. Importantly the present study had greater power to explore these associations due to higher number of genetic instruments for each HSM (apart from HSM5) as compared to the alpha-angle study which had only 8 genetic instruments (12). Interestingly, in the alpha-angle study there was strong evidence that a genetic predisposition to hip osteoarthritis caused an increase in alpha-angle (i.e. cam morphology) and in the present study the same was seen with HSM3 which resembles a cam-type hip with a bulging of the lateral aspect of the femoral head. This adds evidence to the hypothesis that in part osteoarthritis results from a recapitulation of dysregulated growth (33). The shared genetic loci and correlations between hip shape and hip osteoarthritis considered alongside these MR findings suggest that other hip shapes such as reduced or increased acetabular coverage represented by HSM1&2 (i.e. acetabular dysplasia and pincer morphology) might have shared underlying genetics with hip osteoarthritis but do not appear to cause hip osteoarthritis.

Our results suggest that, at least within the context of the older adult population studied here, changes in hip shape are unlikely to play a causal role in the development of hip osteoarthritis. These findings have implications for treatments targeting hip shape with a view to delaying onset or slowing the progression of hip osteoarthritis in older adults including surgery that are currently being investigated in randomised trials (8, 34). On the other hand, rare extremes of hip shape such as developmental dysplasia of the hip which presents in symptomatic children are not well represented in our study population. This is because individuals with such conditions typically undergo hip replacements before the age of 40 years old, making them ineligible for inclusion in the imaging study. Moreover, several rare genetic variants causing more severe forms of hip shape have been identified (35), which are unlikely to have been well proxied by genetic instruments derived from our hip shape GWAS.

To understand if the HSMs were appropriately instrumented by the GWAS results, we also conducted MR analyses between HSMs and hip fracture. These analyses showed strong evidence of a causal association between hip shape and risk of hip fracture. Specifically, higher neck shaft angle (HSM2), wider femoral neck (HSM4) and less acetabular coverage (HSM2&4) were associated with an increased risk of hip fracture. These findings support the observational results seen in UKB and a recent meta-analysis of observational studies, which reported an elevated risk of hip fracture with increased femoral neck shaft angle and femoral neck width (16). These findings also align with those of our recent MR study, which found that increased femoral neck width acts causally to increase risk of hip fracture (36). Taken together, our results suggest that hip shape is a risk factor for hip fracture, supporting the validity and power of these hip shape instruments and adding further weight to the null associations seen for hip osteoarthritis.

This study combined two cohorts which evaluated hip shape using high resolution DXA scans, commonly used to screen for osteoporosis risk. Automatically extracting additional hip shape information from DXA scans in terms of predicting hip osteoarthritis and fracture risk is an attractive proposition. Even if hip shape variance is not causally associated with hip osteoarthritis, these measures could still be important predictive factors. The majority of previous hip shape studies have focused on European populations, but in this study we included an East Asian ancestry group, which broadens the generalisability of our findings (16, 37). However, conducting trans-ancestry GWAS introduces complexity, due to the inherent heterogeneity between populations (38). In this study we used a conservative random-effects model to take account of this heterogeneity and limit the possibility of spurious false positive results driven by the larger size of UKB. Given the disparity in sample size between UKB and SC, we felt it unwise to explore ethnic differences directly in this study. That said, it is known that variations in hip shape exist between European and East Asian populations (17). Future work, harnessing the resources of these large population studies with genetic data, could explore the incorporation of polygenic risk scores to assess whether their inclusion can enhance the performance of disease prediction models (39). In addition, if more cohorts become available then superior trans-ancestry GWAS techniques become possible such as meta-regression of multi-ancestry genetic association (MR-MEGA) (40).

A limitation of this study is that the average age of the participants was ∼60 years old, potentially leading to the coexistence of osteoarthritis in some individuals. However, the majority of participants did not exhibit any signs of radiological osteoarthritis (∼90%), and all outline points, which were placed around the contour of the bones, were manually checked to make sure they did not encompass osteophytes (41). Even if osteophytes were inadvertently included in our measures of hip shape, one would expect this to lead to false positive results from MR rather than the null associations found.

In conclusion, we present findings of a GWAS that investigated hip shape variation in the form of 10 HSMs. Despite finding considerable genetic overlap between hip shape and hip osteoarthritis, causal analyses suggest that rather than causing hip osteoarthritis, changes in hip shape could develop either in tandem with or in response to hip osteoarthritis. On the other hand, there was evidence that hip shape is a causal risk factor for hip fracture. Finally, these results raise doubts over the likely effectiveness of interventions in older individuals intended to reduce hip osteoarthritis progression through modification of hip shape.

## Supporting information

Supplementary Figures and Tables are included

## Acknowledgements

The authors would like to thank the Musculoskeletal Research Unit patient and public involvement group at the University of Bristol for their input into planning our research. This work has been conducted using the UK Biobank resource (application number 17295).

## Funding and grant award information

BGF is supported by a NIHR Academic Clinical Lectureship and was previously supported by a Medical Research Council (MRC) Clinical Research Training Fellowship (MR/S021280/1). RE, MF, FS were supported, and this work is funded by a Wellcome Trust collaborative award (209233/Z/17/Z). MF conducted this work whilst working at the University of Bristol but is now employed by Boehringer Ingleheim UK and Ireland. CL is funded by a Sir Henry Dale Fellowship jointly funded by the Wellcome Trust and the Royal Society (223267/Z/21/Z). This research was funded in whole, or in part, by the Wellcome Trust [Grant numbers 080280/Z/06/Z, 20378/Z/16/Z, 223267/Z/21/Z]. For the purpose of open access, the authors have applied a CC BY public copyright licence to any Author Accepted Manuscript version arising from this submission. NCH acknowledges support from the Medical Research Council (MRC) [MC_PC_21003; MC_PC_21001] and National Institute for Health and Care Research (NIHR) Southampton Biomedical Research Centre, University of Southampton and University Hospital Southampton NHS Foundation Trust, Southampton, UK. JPK is funded by a National Health and Medical Research Council (Australia) Investigator grant (GNT1177938). SW is supported by the “Strategic Priority Research Program” of the Chinese Academy of Sciences (Grant No. XDB38020400) and Shanghai Municipal Science and Technology Major Project, Grant No.2017SHZDZX01. XG is supported by Shanghai Municipal Science and Technology Major Project (Grant No. 2017SHZDZX01).

## Data availability statement

The HSM GWAS meta-analysis summary statistics will be uploaded to the GWAS catalog (https://www.ebi.ac.uk/gwas/). The individual level data from this study concerning UKB participants is available via their data showcase. Users must be registered with UK Biobank to access their resources (https://bbams.ndph.ox.ac.uk/ams/).

## References

1. Agricola R, Heijboer MP, Bierma-Zeinstra SMA, Verhaar JAN, Weinans H, Waarsing JH. Cam impingement causes osteoarthritis of the hip: a nationwide prospective cohort study (CHECK). Annals of the Rheumatic Diseases. 2013;72(6):918–23.

2. Ganz R, Parvizi J, Beck M, Leunig M, Nötzli H, Siebenrock KA. Femoroacetabular Impingement: A Cause for Osteoarthritis of the Hip. Clinical Orthopaedics and Related Research. 2003;417:112–20.

3. Faber BG, Frysz M, Tobias JH. Unpicking observational relationships between hip shape and osteoarthritis: hype or hope? Curr Opin Rheumatol. 2020;32(1):110–8.

4. Lee H, Aronson J. Association or causation? How do we ever know? [Available from: https://catalogofbias.org/2019/03/05/association-or-causation-how-do-we-ever-know/.

5. van Klij P, Heijboer MP, Ginai AZ, Verhaar JAN, Waarsing JH, Agricola R. Cam morphology in young male football players mostly develops before proximal femoral growth plate closure: a prospective study with 5-yearfollow-up. Br J Sports Med. 2019;53(9):532–8.

6. van Klij P, Heerey J, Waarsing JH, Agricola R. The Prevalence of Cam and Pincer Morphology and Its Association With Development of Hip Osteoarthritis. J Orthop Sports Phys Ther. 2018;48(4):230–8.

7. Murphy NJ, Eyles JP, Hunter DJ. Hip Osteoarthritis: Etiopathogenesis and Implications for Management. Adv Ther. 2016;33(11):1921–46.

8. Palmer AJR, Ayyar Gupta V, Fernquest S, Rombach I, Dutton SJ, Mansour R, et al. Arthroscopic hip surgery compared with physiotherapy and activity modification for the treatment of symptomatic femoroacetabular impingement: multicentre randomised controlled trial. BMJ. 2019;364:l185.

9. Davies NM, Holmes MV, Davey Smith G. Reading Mendelian randomisation studies: a guide, glossary, and checklist for clinicians. BMJ. 2018;362:k601.

10. Paternoster L, Tilling K, Davey Smith G. Genetic epidemiology and Mendelian randomization for informing disease therapeutics: Conceptual and methodological challenges. PLoS Genet. 2017;13(10):e1006944.

11. Sanderson E, Glymour MM, Holmes MV, Kang H, Morrison J, Munafò MR, et al. Mendelian randomization. Nature Reviews Methods Primers. 2022;2(1):6.

12. Faber BG, Frysz M, Hartley AE, Ebsim R, Boer CG, Saunders FR, et al. A Genome-Wide Association Study Meta-Analysis of Alpha Angle Suggests Cam-Type Morphology May Be a Specific Feature of Hip Osteoarthritis in Older Adults. Arthritis Rheumatol. 2023;75(6):900–9.

13. Faber BG, Baird D, Gregson CL, Gregory JS, Barr RJ, Aspden RM, et al. DXA-derived hip shape is related to osteoarthritis: findings from in the MrOS cohort. Osteoarthritis Cartilage. 2017;25(12):2031–8.

14. Gregory JS, Testi D, Stewart A, Undrill PE, Reid DM, Aspden RM. A method for assessment of the shape of the proximal femur and its relationship to osteoporotic hip fracture. Osteoporos Int. 2004;15(1):5–11.

15. Frysz M, Faber BG, Ebsim R, Saunders FR, Lindner C, Gregory JS, et al. Machine-learning derived acetabular dysplasia and cam morphology are features of severe hip osteoarthritis: findings from UK Biobank. J Bone Miner Res. 2022.

16. Fajar JK, Taufan T, Syarif M, Azharuddin A. Hip geometry and femoral neck fractures: A meta-analysis. J Orthop Translat. 2018;13:1–6.

17. Zheng J, Frysz M, Faber BG, Lin H, Ebsim R, Ge J, et al. Comparison between UK Biobank and Shanghai Changfeng suggests distinct hip morphology may contribute to ethnic differences in the prevalence of hip osteoarthritis. Osteoarthritis Cartilage. 2023.

18. Rangamaran VR, Uppili B, Gopal D, Ramalingam K. EasyQC: Tool with Interactive User Interface for Efficient Next-Generation Sequencing Data Quality Control. J Comput Biol. 2018;25(12):1301–11.

19. Hemani G. Random-METAL 2022 [updated 8/8/2022. Available from: https://zenodo.org/badge/latestdoi/91003955.

20. Willer CJ, Li Y, Abecasis GR. METAL: fast and efficient meta-analysis of genomewide association scans. Bioinformatics. 2010;26(17):2190–1.

21. Hemani G, Tilling K, Davey Smith G. Orienting the causal relationship between imprecisely measured traits using GWAS summary data. PLoS Genet. 2017;13(11):e1007081.

22. Nethander M, Coward E, Reimann E, Grahnemo L, Gabrielsen ME, Wibom C, et al. Assessment of the genetic and clinical determinants of hip fracture risk: Genome-wide association and Mendelian randomization study. Cell Rep Med. 2022;3(10):100776.

23. Burgess S, Davies NM, Thompson SG. Bias due to participant overlap in two-sample Mendelian randomization. Genet Epidemiol. 2016;40(7):597–608.

24. Baird DA, Evans DS, Kamanu FK, Gregory JS, Saunders FR, Giuraniuc CV, et al. Identification of Novel Loci Associated With Hip Shape: A Meta-Analysis of Genomewide Association Studies. J Bone Miner Res. 2019;34(2):241–51.

25. Boer CG, Hatzikotoulas K, Southam L, Stefansdottir L, Zhang Y, Coutinho de Almeida R, et al. Deciphering osteoarthritis genetics across 826,690 individuals from 9 populations. Cell. 2021;184(18):4784–818 e17.

26. Lindner C, Thiagarajah S, Wilkinson JM, Panoutsopoulou K, Day-Williams AG, Cootes TF, et al. Investigation of association between hip osteoarthritis susceptibility loci and radiographic proximal femur shape. Arthritis Rheumatol. 2015;67(8):2076–84.

27. Faber BG, Ebsim R, Saunders FR, Frysz M, Gregory JS, Aspden RM, et al. Cam morphology but neither acetabular dysplasia nor pincer morphology is associated with osteophytosis throughout the hip: findings from a cross-sectional study in UK Biobank. Osteoarthritis Cartilage. 2021;29(11):1521–9.

28. Heppenstall SV, Ebsim R, Saunders FR, Lindner C, Gregory JS, Aspden RM, et al. Hip geometric parameters are associated with radiographic and clinical hip osteoarthritis: findings from a cross-sectional study in UK Biobank. Osteoarthritis Cartilage. 2023.

29. Bischoff-Ferrari HA. The role of falls in fracture prediction. Curr Osteoporos Rep. 2011;9(3):116–21.

30. Valdes AM, Spector TD, Tamm A, Kisand K, Doherty SA, Dennison EM, et al. Genetic variation in the SMAD3 gene is associated with hip and knee osteoarthritis. Arthritis Rheum. 2010;62(8):2347–52.

31. Zhai G, Dore J, Rahman P. TGF-beta signal transduction pathways and osteoarthritis. Rheumatol Int. 2015;35(8):1283–92.

32. Sorial AK, Hofer IMJ, Tselepi M, Cheung K, Parker E, Deehan DJ, et al. Multi-tissue epigenetic analysis of the osteoarthritis susceptibility locus mapping to the plectin gene PLEC. Osteoarthritis Cartilage. 2020;28(11):1448–58.

33. Aspden RM. Osteoarthritis: a problem of growth not decay? Rheumatology (Oxford). 2008;47(10):1452–60.

34. Griffin DR, Dickenson EJ, Wall PDH, Achana F, Donovan JL, Griffin J, et al. Hip arthroscopy versus best conservative care for the treatment of femoroacetabular impingement syndrome (UK FASHIoN): a multicentre randomised controlled trial. Lancet. 2018;391(10136):2225–35.

35. Zamborsky R, Kokavec M, Harsanyi S, Attia D, Danisovic L. Developmental Dysplasia of Hip: Perspectives in Genetic Screening. Med Sci (Basel). 2019;7(4).

36. Tobias JH, Nethander M, Faber BG, Heppenstall SV, Ebsim R, Cootes T, et al. FEMORAL NECK WIDTH GENETIC RISK SCORE IS A NOVEL INDEPENDENT RISK FACTOR FOR HIP FRACTURES. Journal of Bone and Mineral Research. 2024;In-Press.

37. Gregory JS, Aspden RM. Femoral geometry as a risk factor for osteoporotic hip fracture in men and women. Med Eng Phys. 2008;30(10):1275–86.

38. Zhang M, Wu S, Du S, Qian W, Chen J, Qiao L, et al. Genetic variants underlying differences in facial morphology in East Asian and European populations. Nat Genet. 2022;54(4):403–11.

39. Sedaghati-Khayat B, Boer CG, Runhaar J, Bierma-Zeinstra SMA, Broer L, Ikram MA, et al. Risk Assessment for Hip and Knee Osteoarthritis Using Polygenic Risk Scores. Arthritis Rheumatol. 2022;74(9):1488–96.

40. Magi R, Horikoshi M, Sofer T, Mahajan A, Kitajima H, Franceschini N, et al. Trans-ethnic meta-regression of genome-wide association studies accounting for ancestry increases power for discovery and improves fine-mapping resolution. Hum Mol Genet. 2017;26(18):3639–50.

41. Faber BG, Ebsim R, Saunders FR, Frysz M, Lindner C, Gregory JS, et al. A novel semi-automated classifier of hip osteoarthritis on DXA images shows expected relationships with clinical outcomes in UK Biobank. Rheumatology (Oxford). 2021.

